# LONG COVID RECOVERY AND EXERCISE ADHERENCE: 32-MONTH STUDY

**DOI:** 10.1101/2025.11.11.25340028

**Authors:** Ana Rolo-Duarte, Daniela Prada, Ana S. M. Carvalho, Ana Borges, Paulo J. G. Bettencourt

**Affiliations:** Department of Physical Medicine and Rehabilitation, Hospital Beatriz Ângelo – ULS Loures/Odivelas, Portugal; Faculty of Medicine, Universidade Católica Portuguesa, Lisboa, Portugal; Católica Research Centre for Psychological, Family and Social Wellbeing (CRC-W), Universidade Católica Portuguesa, Lisboa, Portugal; Center for Interdisciplinary Research in Health, Universidade Católica Portuguesa, Lisboa, Portugal

**Author notes:** **Corresponding author**. Ana Rolo-Duarte. Hospital Beatriz Ângelo. Av. Carlos Teixeira 3, 2674-514 Loures, Portugal.

**Keywords:** Long COVID, Rehabilitation, Patient Compliance, Exercise Therapy

## Abstract

**Objective:** To evaluate symptom progression in COVID-19 survivors, adherence to prescribed exercise therapy, and its association with pre-infection physical activity at 21 days (T0), 6 months (T1), and 32 months (T2) post-discharge.

**Design:** Retrospective longitudinal study in a hospital-based rehabilitation unit in Portugal. The cohort included 276 patients (mean age 56.6 ± 13.5 years) with confirmed SARS-CoV-2 infection.

**Results:** Adherence was higher among patients reporting prior physical activity (48.8%; p = .003). Symptom prevalence declined over time: dyspnea (T0 = 22.4%, T2 = 7.3%), fatigue (T0 = 32.4%, T2 = 14.5%), and pain (T0 = 17.6%, T2 = 4.8%). Asymptomatic cases increased from 27.4% (T0) to 54.5% (T2). Early adherence, particularly by day 15, was associated with continued participation at day 21, and adherence at day 21 correlated with reduced dyspnea at follow-up (p = .02). Importantly, patients who remained symptomatic at day 21 took significantly longer to recover (t = –6.386; p < .001), indicating this time point as a prognostic marker of delayed resolution of exercise-modifiable symptoms.

**Conclusion:** Early initiation of individualized, structured exercise proved safe, adaptable, and associated with reduced symptom burden, especially dyspnea. Persistence of symptoms at day 21 highlights the prognostic value of early follow-up and underscores the decisive role of timely rehabilitation engagement. Structured home-based and tele-rehabilitation programs supported adherence and accessibility, reinforcing exercise as a cornerstone of long COVID management and potentially applicable to other post-respiratory rehabilitation contexts.

## INTRODUTION

COVID-19 is a multisystemic disease with heterogeneous clinical presentations. While many individuals experience mild and self-limiting illness, the pandemic caused an estimated 15.9 million deaths globally in 2020–2021.[1] Although the World Health Organization (WHO) declared its end in May 2023, the long-term health consequences of SARS-CoV-2 infection remain a major concern.[2]

Long COVID, defined by the WHO as the persistence or emergence of symptoms three months after infection, lasting at least two months without alternative explanation, affects up to 50% of hospitalized patients [3] and as many as 70% of those with severe disease.[4] Fatigue, dyspnea, and cognitive impairment are among the most frequent sequelae, contributing to functional limitations, reduced quality of life, and delayed return to work.[5]

Multiple mechanisms have been implicated in its pathophysiology, including organ damage, chronic inflammation, endothelial dysfunction, microthrombosis, and autoimmunity.[6–9] Musculoskeletal impairment and critical illness myopathy, already well recognized before the pandemic,[10] are now acknowledged as contributors to long COVID, aggravating weakness and exercise intolerance.[8–10]

Physical inactivity is associated with worse COVID-19 outcomes, whereas higher cardiorespiratory fitness appears protective.[11] Individualized, low-to moderate-intensity exercise has been recommended to support recovery and improve function,[12] and phase-adapted rehabilitation strategies have been proposed.[13] However, uncertainties remain regarding the optimal exercise modality, intensity, and particularly the timing of intervention.[14]]

This study evaluated symptom progression and adherence to prescribed exercise therapy in COVID-19 survivors over 32 months. Special attention was given to the prognostic significance of day 21 post-discharge, to determine whether early rehabilitation engagement influences recovery trajectories and long-term outcomes

## METHODS

### Study Design and Ethical Approval

During the COVID-19 pandemic, to better characterize the clinical course and ensure continuity of Physical Medicine and Rehabilitation (PM&R) care initiated during hospitalization at Hospital Beatriz Ângelo (HBA), Portugal, patients were systematically evaluated in the hospital’s outpatient PM&R clinic following discharge. Informed consent was obtained by phone and documented in the medical records, approved by the Ethics Committee. The study was approved by the research department and the Ethics Committee of HBA, with Ethics Approval Reference 3734/2021_MJHMAB/FB and 4692/2025_MJH/CL for study n.°576_LH n.°329. The data were collected between 1 May 2020 and 31 July 2025. Data were accessed for research purposes on the 31 July 2025. Only the medical authors who were responsible for monitoring the patients had access to the participants’ identities.

### Participants

Adult patients with clinically and laboratory-confirmed SARS-CoV-2 infection, diagnosed by reverse transcription polymerase chain reaction (RT-PCR), were included.

Patients were considered eligible after achieving clinical stability in the general COVID-19 ward. In-hospital exercise therapy was initiated in those meeting predefined clinical safety criteria, including comprehensive assessment of respiratory, cardiovascular, and neurological systems, as well as hematologic and laboratory parameters [15,16] (see S1 Appendix).

The final sample included 276 hospitalized patients, aged 20 to 86 years (mean = 56.6 ± 13.5), of whom 194 (71.1%) were male. Among them, 163 patients (59.7%) required admission to the intensive care unit (ICU), mainly due to hypoxemic respiratory failure. Invasive mechanical ventilation (IMV) was necessary in 79 patients (29.2%), and prone positioning was frequently employed.Prior physical activity was reported by 97 patients (35.1%) and was defined as meeting the World Health Organization’s recommended levels: 150–300 minutes of moderate-intensity or 75–150 minutes of vigorous-intensity aerobic activity per week.[17]

The most common comorbidities were hypertension (58.3%), obesity (57.4%), dyslipidemia (41.1%), and diabetes mellitus (32.4%). Other relevant conditions included psychiatric disorders (10.2%), malignancy (7.0%), chronic pulmonary disease (6.6%), and asthma (6.1%). Notably, 45.5% of patients had no comorbidities.

### In-Hospital Rehabilitation Program

During isolation, the PM&R department developed an exercise plan to continue rehabilitation started during hospitalization. After clinical stabilization (typically by day 2–3), the team provided education and individualized instruction.

Patients received a printed guide with 10 standardized exercises (breathing, limb and trunk strength, and balance), illustrated with photos, plus a resistance band (TheraBand®). Intensity was progressively adapted (3–20 repetitions), with adjustments based on tolerance under physiotherapist supervision. Oxygen saturation and heart rate were monitored with a pulse oximeter, and symptoms were assessed using the Modified Borg Scale (mBorg). Exercise was discontinued if saturation dropped >4% from baseline, fell below 90%, or if fatigue, dyspnea, or pain exceeded 3 on the mBorg. Progression, including increased repetitions, was implemented under physiotherapist supervision.

### Post-Discharge Home-Based Program

Upon discharge, all instructional materials and equipment except for the pulse oximeter were provided to patients to continue the home-based program. Exercise compliance was monitored through structured follow-up, including three telephone contacts by the clinical team, forming part of a tele-rehabilitation strategy to support adherence and address any emerging complications related to the prescribed exercise plan. The first call occurred between the 5th and 10th day after discharge, the second on the 15th day (both by physiotherapists), and the third on the 21st day, conducted by a physician. Patients were also encouraged to begin walking after the isolation period, with appropriate safety precautions (Figure 1).

**Figure 1.**
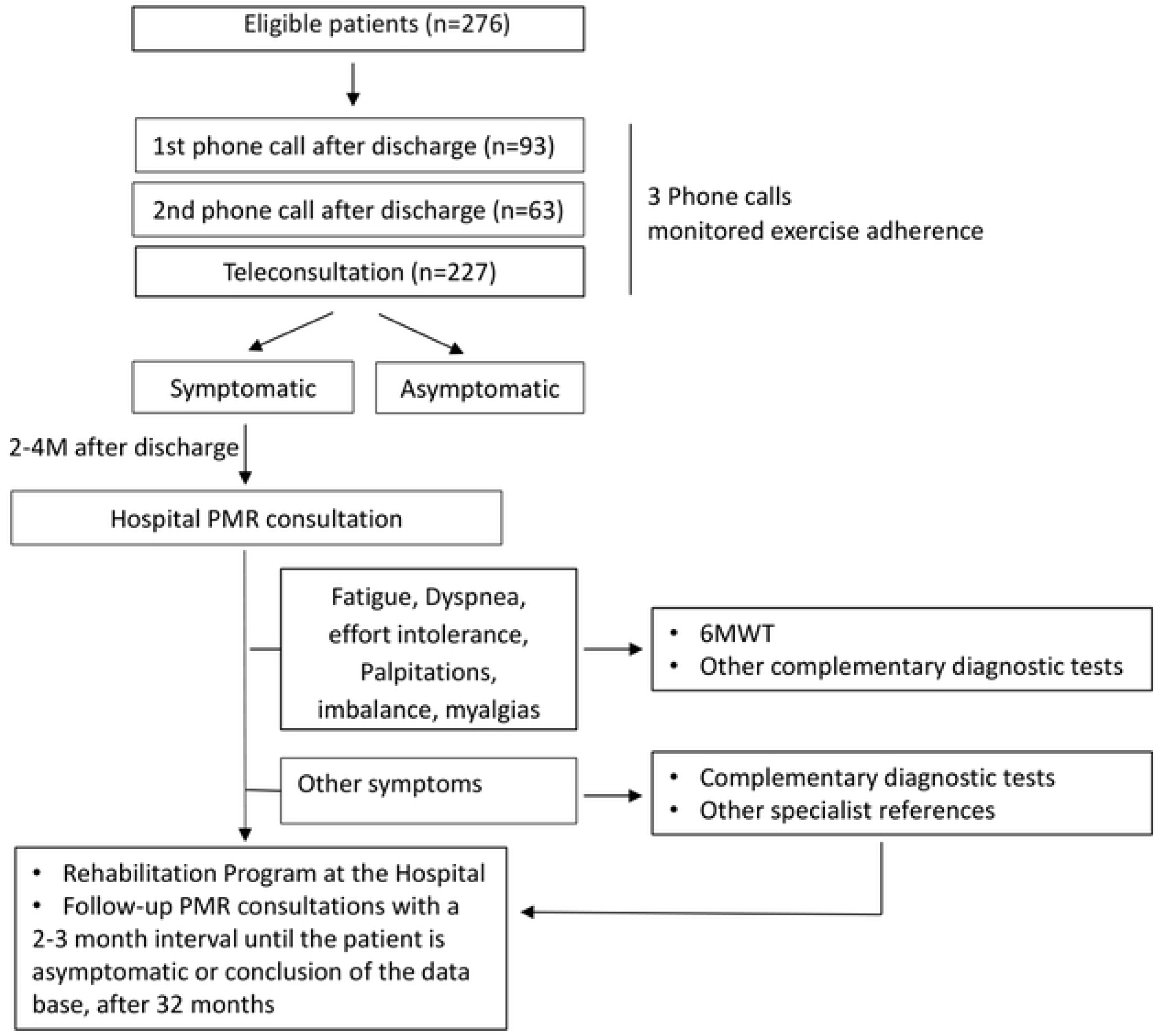
Flow chart of the study

Due to fluctuations in healthcare response capacity during the pandemic, only 231 of the 276 patients (83.7%) received at least one follow-up call. Due to the absence of adherence data, the remaining 45 patients (16.3%) could not be included in analyses examining exercise compliance or symptom progression related to follow-up timing. Of those contacted, 93 (33.7%) received the first call, 63 (22.8%) received the second, and 227 (82.2%) were reached on the 21st day post-discharge.

### Outpatient Follow-Up and Clinical Monitoring

Compliance with the prescribed exercise program was assessed, and symptoms were recorded during the day-21 teleconsultation. If symptoms or disability persisted, a face-to-face PM&R consultation was scheduled between the second and fourth months, post-discharge, depending on appointment availability.

Pain was defined as new-onset or worsening of prior symptoms. When appropriate, a 6-minute walk test (6MWT) was performed during in-person visits to assess oxygen desaturation, heart rate, and functional capacity. Referrals to other specialties and neuropsychology were made when needed.[18] Follow-up diagnostics were limited due to infection risk and logistical constraints.

### Rehabilitation Prescription and Long-Term Monitoring

During in-person PM&R consultations, patients received a comprehensive clinical assessment and a tailored rehabilitation prescription (RP), which included physiotherapy and occupational therapy, primarily delivered at the hospital. Follow-up appointments were scheduled every 2 to 3 months. Monitoring continued until rehabilitation-responsive symptoms (e.g., fatigue, dyspnea, pain, myalgias, peripheral sequelae, muscle weakness, imbalance, reduced effort tolerance) had resolved, or until the study database closed at 32 months.

### Study Variables

Variables included demographic data (age, sex), ICU admission, IMV, discharge date, prior physical activity, comorbidities, follow-up contact dates, exercise adherence, and symptom evolution (see S2 Appendix).

### Data Analysis

Statistical analysis was performed using **IBM** Statistical Package for the Social Sciences (SPSS), version 28 (IBM Corp., Armonk, NY, USA). Descriptive and frequency analyses were conducted for demographic and clinical variables. Frequency analyses, chi-square tests of independence, and independent-sample t-tests were used to assess associations between the study variables. Patients who did not receive any follow-up calls (n = 45;16.3%) were excluded from analyses involving exercise adherence and its association with outcomes, due to lack of relevant data. However, they were retained in overall descriptive statistics where applicable. No data imputation was performed. Full details of the statistical procedures are provided in S3 Appendix.

## RESULTS

### Exercise adherence

Most patients adhered consistently to the prescribed exercise program throughout follow-up (Figure 2). A significant association was found between adherence at day 15 and adherence at day 21 post-discharge (Table 1; χ² = 4.712; p = .03), with 83.6% of individuals who exercised on day 15 also adhering on day 21 (Table 1).

**Figure 2.**
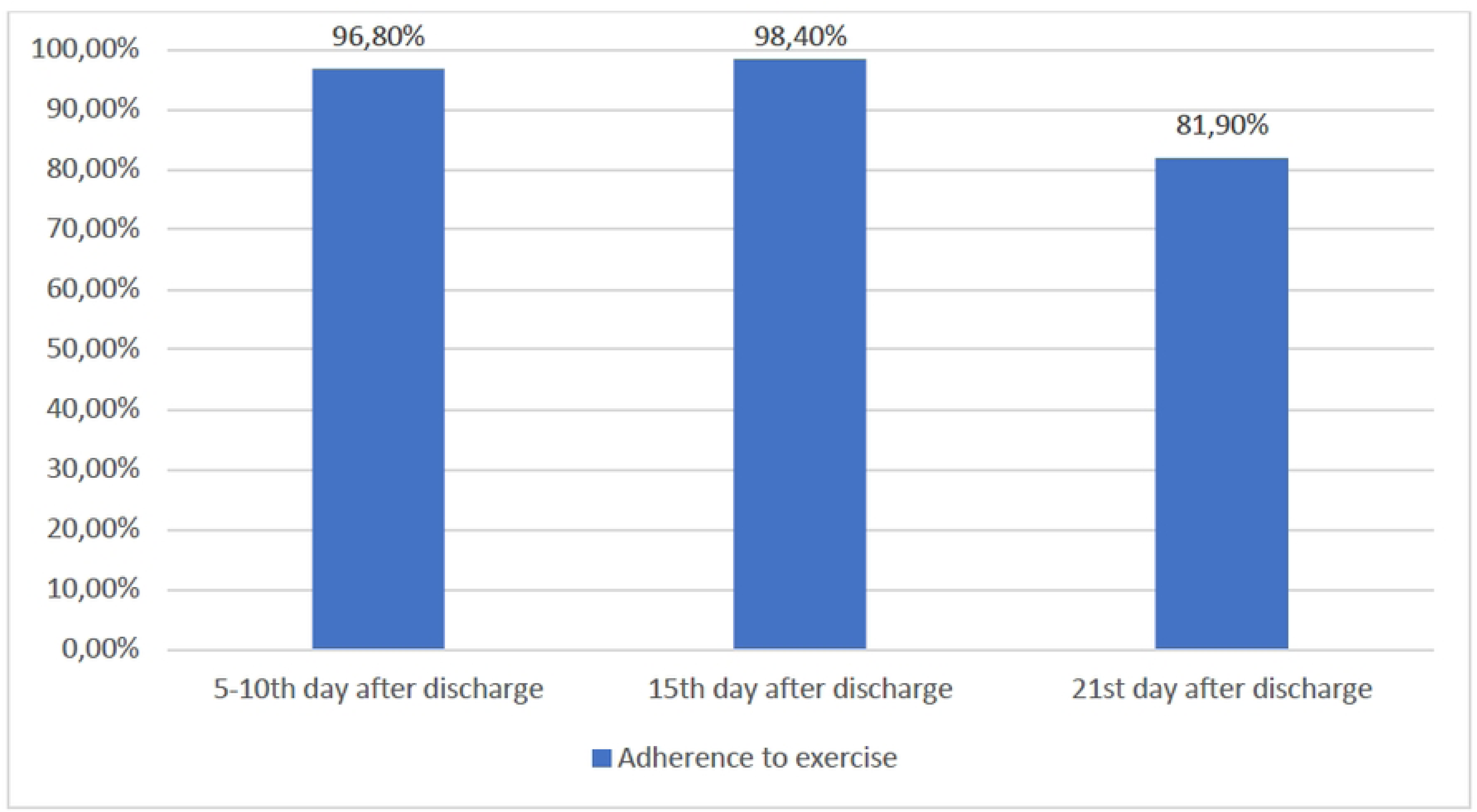
*Adherence to exercise at three follow-up points*

**Table 1.** Association between early exercise adherence and day 21 adherence in covid-19 survivors.

| Adherence to exercise | | | | 21st day after discharge | | $\chi^2$ |
| --- | --- | --- | --- | --- | --- | --- |
|  |  |  |  | No | Yes |  |
| 5-10th | day | after | No | 1 (33.3%) | 2 (66.7%) | 1.552 ($p=.21$) |
| discharge |  |  | Yes | 9 (10.3%) | 78 (89.7%) |  |
| 15th | day | after | No | 1(100.0%)* | 0 (0.0%)* | 4.712 ($p=.03$) |
| discharge |  |  | Yes | 10(16.4%)* | 51 (83.6%)* |  |

\* $p < .05$

### Symptom Burden and Adherence

Intervention strategies aimed at enhancing patients’ understanding of the importance of exercise for physical rehabilitation promoted sustained adherence, even 21 days post-discharge. Additionally, analysis of long COVID symptoms revealed that the majority (83.2%) of patients who performed the exercises on day 21 did not experience dyspnea post-discharge (𝜒2(1) = 5.289, p = .02) (Table 2). The average follow-up time in the PM&R consultation was 6.1 months (M = 6.1; SD = 6.2).

**Table 2.** Day 21 Exercise and Post-COVID Symptom Burden.

| Symptoms | Adherence to exercise | | | $\chi^2$ |
| --- | --- | --- | --- | --- |
|  |  | No | Yes |  |
| Pain | No | 32 (18.5%) | 141 (81.5%) | 0.193 ($p=.66$) |
|  | Yes | 8 (21.6%) | 29 (78.4%) |  |
| Fatigue | No | 28 (17.5%) | 132 (82.5%) | 0.915 ($p=.34$) |
|  | Yes | 12 (23.5%) | 39 (76.5%) |  |
| Dyspnea | No | 31 (16.8%)* | 154 (83.2%)* | 5.289 ($p=.02$) |
|  | Yes | 9 (36.0%)* | 16 (64.0%)* |  |
| Neurologic sequelae | No | 39 (19.3%) | 163 (80.7%) | 0.231 ($p=.63$) |
|  | Yes | 1 (12.5%) | 7 (87.5%) |  |
| Myalgias | No | 37 (18.6%) | 162 (81.4%) | 0.509 ($p=.48$) |
|  | Yes | 3 (27.3%) | 8 (72.7%) |  |
| Decline in muscular strength | No | 35 (18.7%) | 152 (81.3%) | 0.121 ($p=.73$) |
|  | Yes | 5 (21.7%) | 18 (78.3%) |  |
| Imbalance | No | 37 (18.5%) | 163 (81.5%) | 0.817 ($p=.37$) |
|  | Yes | 3 (30.0%) | 7 (70.0%) |  |
| Decreased effort tolerance | No | 40 (19.5%) | 165 (80.5%) | 1.205 ($p=.27$) |
|  | Yes | 0 (0.0%) | 5 (100.0%) |  |
| Paresthesias | No | 39 (19.0%) | 166 (81.0%) | 0.003 ($p=.96$) |
|  | Yes | 1 (20.0%) | 4 (80.0%) |  |

\* $p<.05$

### Effect of Prior Physical Activity on Adherence

Patients were asked about their exercise habits prior to COVID-19 infection. A significant association was observed between previous habits and adherence on day 21 (𝜒2(1) = 8.991, p = .003), Nearly half of adherent patients (48.8%) reported meeting WHO physical activity guidelines prior to COVID-19 infection. (Table 3).

**Table 3.**
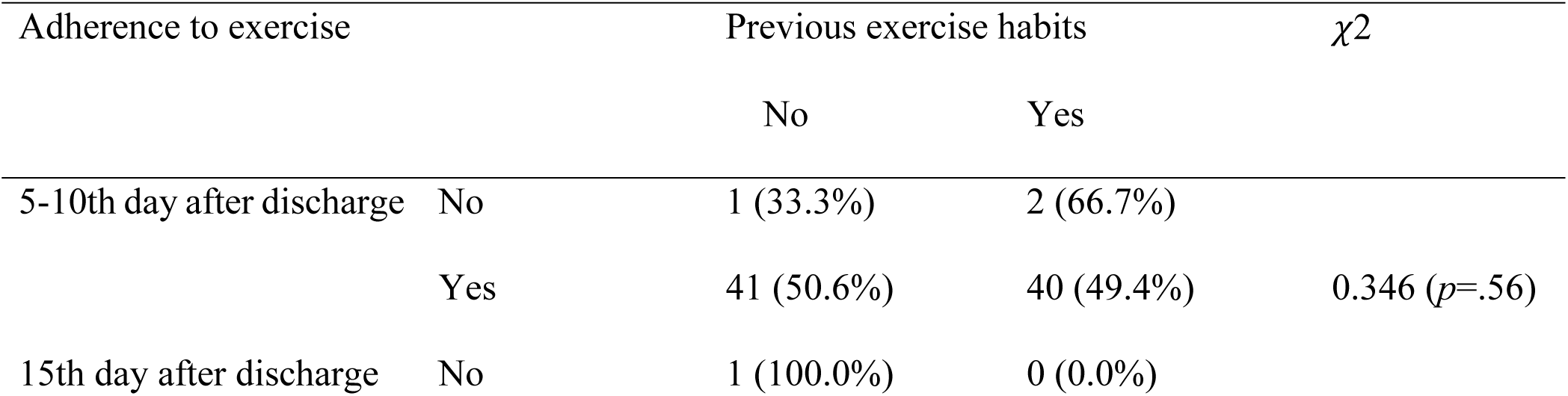
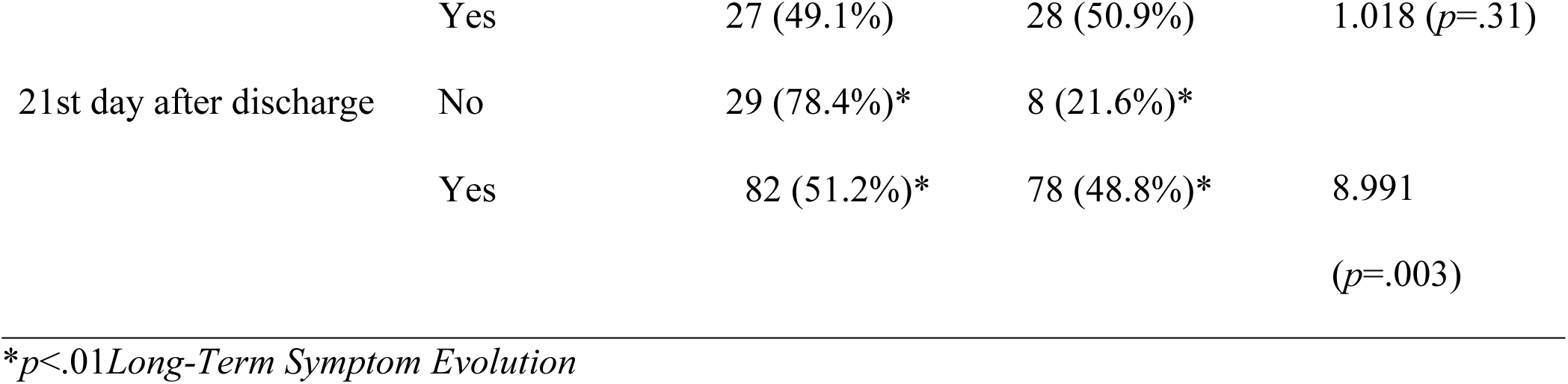
Pre-Infection Physical Activity and Day-21 Exercise Adherence.

Most patients were aged 55–64 years (34.4%, 50.0%, and 28.0% at T0, T1, and T2, respectively). Males predominated at T0 (65.6%) and T2 (72.0%), while females were more frequent at T1 (58.3%). Symptoms were assessed at follow-up consultations every 2–3 months, with data compared at T0 (21 days), T1 (6 months), and T2 (32 months) post-discharge. Figure 3 shows symptom evolution across these time points.

**Figure 3.**
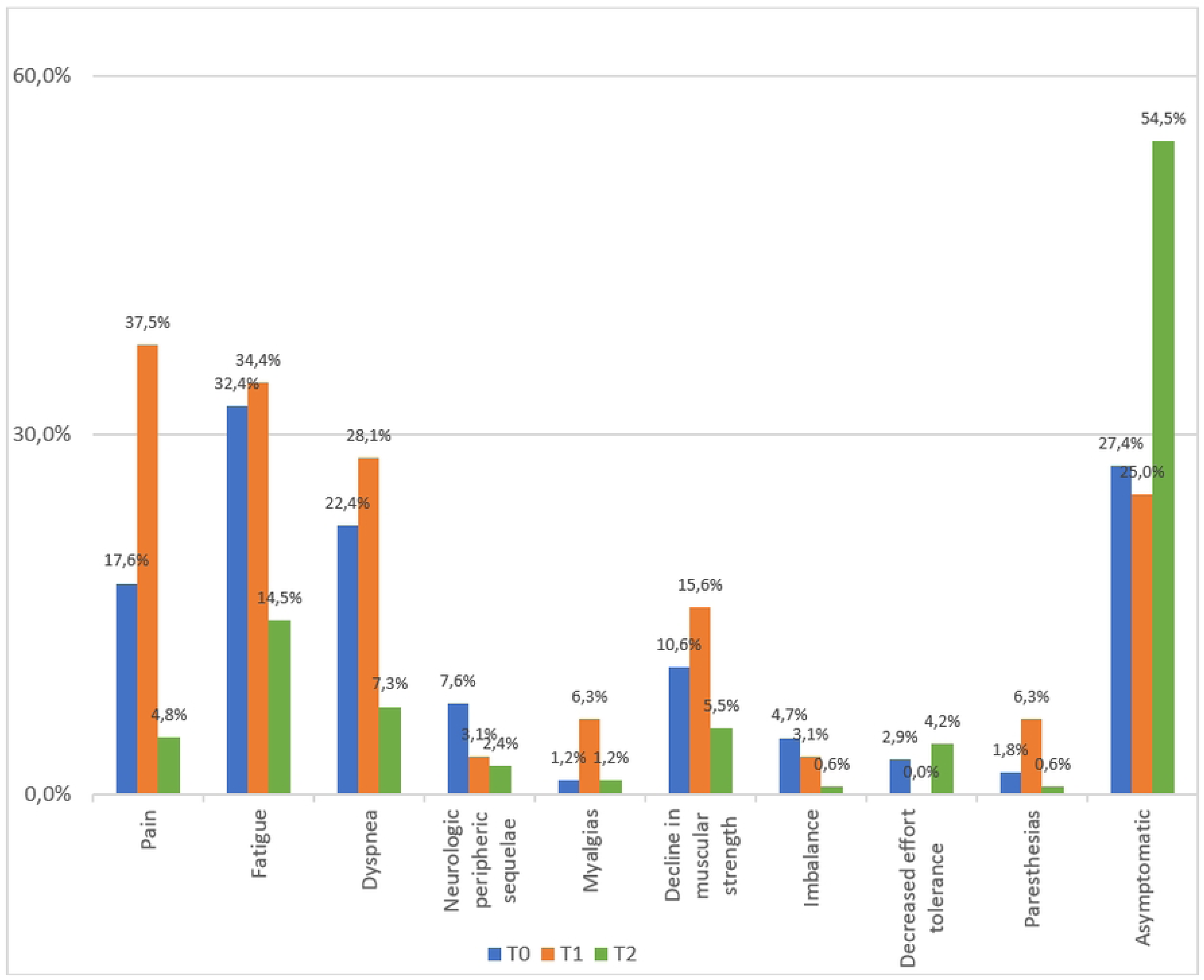
*Evolution of the most frequent post-COVID symptoms at 21 days (T0), 6 months (T1), and 32 months (T2) after discharge*

At T0, the most common symptoms were fatigue (32.4%), dyspnea (22.4%), and pain (17.6%); at T1, pain (37.5%), fatigue (34.4%), and dyspnea (28.1%); and at T2, fatigue (14.5%), dyspnea (7.3%), and reduced strength (5.5%). The proportion of asymptomatic patients increased from 27.4% at T0 to 54.5% at T2.

A significant association was found between symptoms reported on the day-21 phone call and delayed resolution of exercise-modifiable symptoms, with symptomatic patients taking longer to become asymptomatic (t = −6.386; p < .001) (Table 4).

**Table 4.** Impact of Day-21 Symptom Status on Time to Resolution of Exercise-Modifiable Symptoms.

| Time to resolution of exercise-modifiable symptoms |  |  |  |
| --- | --- | --- | --- |
| Symptomatic patients | n | *M (SD)* | *t* |
| No | 73 | 4.32 (3.80) |  |
| Yes | 95 | 9.48 (6.06) | -6.386 ($p<.001$) |

## DISCUSSION

This retrospective longitudinal study highlighted the potential benefits of early, structured exercise therapy in post-COVID rehabilitation and provided novel data on symptom evolution up to 32 months after hospital discharge.

The cohort was predominantly male and middle-aged, with a high prevalence of comorbidities - particularly hypertension, followed by obesity and dyslipidemia. Most participants were aged between 55 and 64 years. In this study, the majority of patients with long COVID were male at T0 and T2, and female at T1. Previous studies identified a higher risk of developing long COVID in association with several factors, including female sex, middle age, obesity, infection severity, hospital admission (particularly requiring oxygen therapy), symptom burden (such as dyspnea and chest pain), and pre-existing comorbidities such as asthma.[19–22] While the age distribution in this cohort aligns with prior studies,[22] the predominance of male patients contrasts with most reports.[20] This discrepancy may reflect the higher proportion of male hospitalizations during the study period. These comorbidities are well-established risk factors for severe COVID-19 and have been linked to delayed recovery and greater symptom burden in post-acute and long COVID populations. In this cohort, early adherence to the exercise regimen, particularly by day 15, was strongly associated with continued participation through day 21, underscoring the value of early rehabilitation engagement. Moreover, patients with a history of physical activity prior to infection demonstrated better adherence post-discharge, reinforcing the influence of lifestyle factors on recovery. Notably, adherence at day 21 was associated with reduced dyspnea at final consultation.

An important finding of this study is the identification of day 21 as a critical prognostic time point. Patients who still reported symptoms and did not adhere to the exercise program at this stage took significantly longer to become asymptomatic. This emphasizes that, while the benefits of exercise in long COVID are recognized, the timing of initiation is decisive: patients who fail to engage in exercise early—within the first three weeks after discharge—are at substantially higher risk of prolonged recovery. Clinically, this suggests that day-21 follow-up is valuable not only for monitoring adherence but also for identifying high-risk patients who may require intensified rehabilitation strategies and closer supervision.

Long-term symptom monitoring revealed a progressive decline in key symptoms (fatigue, dyspnea, and pain) and an increase in asymptomatic individuals over time. However, patients who remained symptomatic at day 21 experienced significantly slower recovery, suggesting early symptom persistence as a potential prognostic marker of prolonged rehabilitation.

The pathophysiological mechanisms underlying long COVID remain incompletely understood. Prolonged systemic inflammatory responses have been proposed as a potential mechanism for long COVID, particularly in patients with mild acute infection who develop persistent symptoms despite no detectable organ damage.[19,23] Several mechanisms have been implicated in the pathogenesis of long COVID, including alterations in the angiotensin-converting enzyme 2 (ACE2) pathway and dysregulation of the renin–angiotensin–aldosterone system (RAAS)[24], endothelial dysfunction [25], neuromuscular impairment26, and autonomic dysregulation.[27] Given these complex mechanisms, interventions that target systemic inflammation and functional impairment—such as exercise—are increasingly supported by evidence. Exercise plays a key role in post-COVID rehabilitation, not only by enhancing functional capacity but also through its anti-inflammatory effects. Regular moderate-intensity exercise reduces systemic inflammatory mediators (CRP, IL-6, TNF-α) through improved immune regulation, increased anti-inflammatory cytokines, and enhanced vagal tone. Given the contribution of chronic low-grade inflammation to long COVID, exercise may directly modulate its pathophysiology and support recovery. [28,29]

This study underscores the importance of individualized rehabilitation strategies and multidisciplinary care in long COVID.30 Exercise interventions were adapted to patient tolerance and adjusted progressively based on symptom response, accommodating the heterogeneity of clinical presentations. Supervision by physiotherapists and physicians, including structured telephone follow-up, enabled safe and remote engagement, particularly during isolation periods. These findings highlight the utility of tele-rehabilitation in supporting adherence and extending access to care, reinforcing its role within comprehensive post-COVID rehabilitation protocols.

Limitations

This study has several limitations. First, pre-existing symptomatology data were unavailable, potentially confounding persistent symptom interpretation. Second, symptom severity assessment was limited by data granularity constraints. Third, excluding patients without follow-up calls may have introduced selection bias. Additionally, this single-center study lacked a control group of patients discharged after non-COVID illnesses. Since prolonged symptoms may occur after community-acquired pneumonia, findings may have broader relevance beyond COVID-19

## CONCLUSION

This study evaluated post-discharge symptoms in COVID-19 survivors and their adherence to a rehabilitation program for long COVID. The findings reinforce the importance of early and individualized rehabilitation, particularly through structured therapeutic exercise. Interventions were safe, adaptable, and associated with reduced symptom burden—most notably dyspnea.

An important finding of this study was the identification of day 21 as a critical prognostic point: patients who remained symptomatic and did not adhere to exercise at this stage took significantly longer to recover. This underscores that early initiation of low- to moderate-intensity exercise under professional supervision is decisive for optimizing recovery, beyond its role as a functional enhancer and immune modulator.

Additionally, the successful implementation of tele-rehabilitation highlights its potential to enhance accessibility and support patient management. Collectively, these results suggest that structured and early rehabilitation strategies may be extended to other populations recovering from severe respiratory illnesses.

## Data Availability

All relevant data supporting the findings of this study are included within the manuscript. The underlying individual-level data contain potentially identifying or sensitive patient information and therefore cannot be made publicly available. De-identified data are available from the corresponding author upon reasonable request and with permission from the Ethics Committee of Hospital Beatriz Ângelo.

## Acknowledgements

The authors would like to acknowledge Gisela Gomes, PT; Patricia Martins, PT; Carla Rodrigues, PT; and Tânia Bastos, PT for performing physiotherapy sessions, conducting follow-up phone calls, and assisting with data collection. The authors also thank Cláudia Abreu, PT for her contribution in designing the patient exercise brochure. Additionally, ARD, DP, and AB acknowledge Ana Filipa Regadas, PT for her support in the organizational aspects of the study.

## Author Disclosures

### Financial Benefits

The authors declare no financial interests or personal gains related to this study.

### Previous Presentation

This research has not been presented in any form, including abstracts, posters, or conferences.

## REFERENCES

1. GBD 2021 Demographics Collaborators. Global age-sex-specific mortality, life expectancy, and population estimates in 204 countries and territories and 811 subnational locations, 1950–2021, and the impact of the COVID-19 pandemic: a comprehensive demographic analysis for the Global Burden of Disease Study 2021. Lancet. 2024;403(10440):1989–2056. doi:10.1016/S0140-6736(24)00476-8

2. World Health Organization. Post COVID-19 condition (Long COVID). Published December 7, 2022. Accessed June 19, 2025. https://www.who.int/europe/news-room/fact-sheets/item/post-covid-19-condition

3. Shi J, Lu R, Tian Y, Wu F, Geng X, Zhai S, et al. Prevalence of and factors associated with long COVID among US adults: a nationwide survey. BMC Public Health. 2025; 25:1758. doi:10.1186/s12889-025-22987-8

4. Davis HE, McCorkell L, Moore Vogel J, Topol EJ. Long COVID: major findings, mechanisms and recommendations. Nat Rev Microbiol. 2023;21(3):133–146. doi:10.1038/s41579-022-00846-2

5. Halpin SJ, McIvor C, Whyatt G, Adams A, Harvey O, McLean L, et al. Postdischarge symptoms and rehabilitation needs in survivors of COVID-19 infection: a cross-sectional evaluation. J Med Virol. 2021;93(2):1013–1022. doi:10.1002/jmv.26368;93(2):1013–1022. doi:10.1002/jmv.26368

6. Xie Y, Bowe B, Al-Aly Z. Long-term cardiovascular outcomes of COVID-19. Nat Med. 2022;28(3):583–590. doi:10.1038/s41591-022-01689-3

7. Carvalho-Schneider C, Laurent E, Lemaignen A, Beaufils E, Bourbao-Tournois C, Laribi S, et al. Follow-up of adults with noncritical COVID-19 two months after symptom onset. Clin Microbiol Infect. 2021;27(2):258–263. doi:10.1016/j.cmi.2020.09.052

8. Agergaard J, Khan BYA, Engell-Sørensen T, Schiøttz-Christensen B, Østergaard L, Hejbøl EK, et al. Myopathy as a cause of Long COVID fatigue: Evidence from quantitative and single fiber EMG and muscle histopathology. Clin Neurophysiol. 2023;148:65–75. doi:10.1016/j.clinph.2023.01.010

9. Ramírez-Vélez R, Legarra-Gorgoñon G, Oscoz-Ochandorena S, García-Alonso Y, García-Alonso N, Oteiza J, et al. Reduced muscle strength in patients with long-COVID-19 syndrome is mediated by limb muscle mass. J Appl Physiol. 2023;134(1):50–58. doi:10.1152/japplphysiol.00599.202

10. Schefold JC, Wollersheim T, Grunow JJ, Luedi MM, Z’Graggen WJ, Weber-Carstens S. Muscular weakness and muscle wasting in the critically ill. J Cachexia Sarcopenia Muscle. 2020;11(6):1399–1412. doi:10.1002/jcsm.12620

11. Rahmati M, Shamsi MM, Khoramipour K, Malakoutinia F, Woo W, Park S, et al. Baseline physical activity is associated with reduced mortality and disease outcomes in COVID-19: A systematic review and meta-analysis. Rev Med Virol. 2022;32(5):e2349. doi:10.1002/rmv.2349

12. Zheng C, Chen XK, Sit CH-P, Liang X, Li MH, Ma ACH-H, et al. Effect of physical exercise-based rehabilitation on long COVID: a systematic review and meta-analysis. Med Sci Sports Exerc. 2024;56(1):143–154. doi:10.1249/MSS.0000000000003280

13. Gutenbrunner C, Nugraha B, Martin LT. Phase-adapted rehabilitation for acute coronavirus disease-19 patients and patients with long-term sequelae of coronavirus disease-19. Am J Phys Med Rehabil. 2021;100(6):533–538. doi:10.1097/PHM.0000000000001762

14. Besnier F, Malo J, Mohammadi H, Clavet S, Klai C, Martin N, et al. Effects of cardiopulmonary rehabilitation on cardiorespiratory fitness and clinical symptom burden in Long COVID: results from the COVID-Rehab randomized controlled trial. Am J Phys Med Rehabil. 2025;104(2):163–171. doi:10.1097/PHM.0000000000002559

15. Zhao HM, Xie YX, Wang C. Recommendations for respiratory rehabilitation in adults with coronavirus disease 2019. Chin Med J (Engl). 2020;133(13):1595–1602. doi:10.1097/CM9.0000000000000848

16. Conceição T, Gonzales AI, Figueiredo F, Vieira DSR, Bundchen DC. Safety criteria to start early mobilization in intensive care units: systematic review. Rev Bras Ter Intensiva. 2017;29(4):509–519. doi:10.5935/0103-507X.20170076

17. Bull FC, Al-Ansari SS, Biddle S, Borodulin K, Buman MP, Cardon G, et al. World Health Organization 2020 guidelines on physical activity and sedentary behaviour. Br J Sports Med. 2020;54(24):1451–1462. doi:10.1136/bjsports-2020-102955

18. Ceravolo MG, Anwar F, Andrenelli E, Udensi C, Qureshi J, Sivan M, et al. Evidence-based position paper on physical and rehabilitation medicine professional practice for persons with COVID-19, including post COVID-19 condition: the European PRM position (UEMS PRM Section). Eur J Phys Rehabil Med. 2023;59(6):789–799. doi:10.23736/S1973-9087.23.08315-6

19. Sudre CH, Murray B, Varsavsky T, Graham MS, Penfold RS, Bowyer RC, et al. Attributes and predictors of long COVID. Nat Med. 2021;27(4):626–631. doi:10.1038/s41591-021-01292-y

20. Moritani I, Yamanaka K, Nakamura T, Tanaka J, Kainuma K, Okamoto M, et al. Prevalence of and risk factors for long COVID following infection with the COVID-19 omicron variant. Med Int (Lond). 2025;5(2):17. doi:10.3892/mi.2025.216

21. Castanares-Zapatero D, Chalon P, Kohn L, Dauvrin M, Detollenaere J, Maertens de Noordhout C, et al. Pathophysiology and mechanism of long COVID: a comprehensive review. Ann Med. 2022;54(1):1473–1487. doi:10.1080/07853890.2022.2076901

22. Guan WJ, Liang WH, Zhao Y, Liang HR, Chen ZS, Li YM, et al. Comorbidity and its impact on 1590 patients with COVID-19 in China: a nationwide analysis. Eur Respir J. 2020;55(5):2000547. doi:10.1183/13993003.00547-2020

23. Wirth KJ, Scheibenbogen C. Dyspnea in post-COVID syndrome following mild acute COVID-19 infections: potential causes and consequences for a therapeutic approach. Medicina (Kaunas). 2022;58(3):419. doi:10.3390/medicina58030419

24. Astin R, Banerjee A, Baker MR, Dani M, Ford E, Hull JH, et al. Long COVID: mechanisms, risk factors and recovery. Exp Physiol. 2023;108(1):12–27. doi:10.1113/EP090802

25. Gleeson M, Bishop NC, Stensel DJ, Lindley MR, Mastana SS, Nimmo MA. The anti-inflammatory effects of exercise: mechanisms and implications for the prevention and treatment of disease. Nat Rev Immunol. 2011;11(9):607–615. doi:10.1038/nri3041

26. Swarnakar R, Yadav SL. Rehabilitation in long COVID-19: A mini-review. World J Methodol. 2022;12(4):235–245. doi:10.5662/wjm.v12.i4.235

27. Vassiliou AG, Vrettou CS, Keskinidou C, Dimopoulou I, Kotanidou A, Orfanos SE. Endotheliopathy in acute COVID-19 and long COVID. Int J Mol Sci. 2023;24(9):8237. doi:10.3390/ijms24098237

28. Jimeno-Almazán A, Pallarés JG, Buendía-Romero Á, Martínez-Cava A, Franco-López F, Sánchez-Alcaraz Martínez BJ, et al. Post-COVID-19 syndrome and the potential benefits of exercise. Int J Environ Res Public Health. 2021;18(10):5329. doi:10.3390/ijerph18105329

29. Gleeson M, Bishop NC, Stensel DJ, Lindley MR, Mastana SS, Nimmo MA. The anti-inflammatory effects of exercise. Nat Rev Immunol. 2011;11(9):607–615. doi:10.1038/nri3041

30. Swarnakar R, Yadav SL. Rehabilitation in long COVID-19: A mini-review. World J Methodol. 2022;12(4):235–245. doi:10.5662/wjm.v12.i4.235

